# Safety and Pharmacokinetics of a Novel Non-Steroidal Mineralocorticoid Receptor Antagonist, KBP-5074, in Hemodialysis and Non-Hemodialysis Patients with Severe Chronic Kidney Disease

**DOI:** 10.1101/2020.05.12.20053314

**Authors:** Jeffrey Connaire, Mark Bush, Jon Ruckle, Fred Yang, Jinrong Liu, Xiaojuan Tan, Ping Wang, Tian Zhou, Min Zhang, Vincent Benn

## Abstract

**Objective:** KBP-5074 is a novel non-steroidal mineralocorticoid receptor antagonist for the potential treatment of hypertension. The purpose of this study was to assess the safety, tolerability, pharmacokinetics, and pharmacodynamics of a single dose of 0.5 mg KBP-5074 in subjects with end stage renal disease on and not on hemodialysis.

**Results:** Single doses of 0.5 mg KBP-5074 were generally well tolerated in both hemodialysis (N=6) and non-hemodialysis (N=5) subjects with severe renal impairment. Of the 11 subjects in the study, there was a single report of hyperkalemia in a hemodialysis subject on Day 14 that was not considered drug-related and was resolved in 2 days. Mean plasma KBP-5074 concentrations peaked at 6 and 4 hours in non-dialysis and dialysis subjects, respectively, and slowly declined through 312 hours postdose. Overall, the KBP-5074 exposures were significantly lower and had a shorter half-life in hemodialysis subjects compared with the non-hemodialysis subjects. However, hemodialysis had minimal direct impact on drug concentrations with negligible drug levels in the dialysate samples. Plasma aldosterone and serum potassium concentrations were generally comparable between non-hemodialysis and hemodialysis subjects, most likely to due to high variability in the hemodialysis subjects.

**Conclusion:** The overall plasma exposure of KBP-5074 was statistically significantly lower in hemodialysis subjects compared to subjects with renal impairment not on hemodialysis.

## 1 Introduction

Hypertension and hypertensive kidney damage pose a significant health risk in the United States and in 2013 were ranked in the top 15 leading causes of morbidity and mortality.^1^ Hypertension is also a major risk factor for other causes of death such as heart failure^2^, stroke, and coronary vascular disease.^3^ Although considerable progress has been made in the treatment of hypertension and renal failure, the development of new therapy remains critical.

Therapies for the treatment of hypertension and hypertensive kidney damage have targeted the mineralocorticoid receptor (MR), which is a member of the steroid receptor family. Mineralocorticoid receptors are found on epithelial cells located in the kidney, colon, salivary, and sweat glands, and promote sodium retention and a loss of potassium and magnesium. Disturbances in these electrolytes may impair cardiac function and increase or enhance the risk of morbid cardiac and vascular events.^4^ Treatments such as aldosterone receptor antagonists have been shown to inhibit the effects of aldosterone signaling at MR, thus preventing sodium retention and loss of potassium.^4^ Mineralocorticoid receptors have also been shown to have a major pathophysiological role in the progression of kidney diseases.^5,6,7^ It has been shown that the inhibition of mineralocorticoid receptor signaling considerably reduces proteinuria in patients with chronic kidney disease (CKD).^5,8,9^

Eplerenone and spironolactone are steroidal MR antagonists (MRAs) that are commonly used for the treatment of hypertension and heart failure in patients with mild to moderate renal impairment; although, their use is limited by the potential risk of hyperkalemia. Therefore, it is critical to develop new MRAs with improved efficacy and lower potential for adverse events (AEs).

KBP-5074 is a novel non-steroidal MRA being developed for the treatment of uncontrolled hypertension and advanced CKD. In *in vitro* studies, KBP-5074 selectively binds to recombinant human MR relative to other recombinant human glucocorticoid, progesterone, or androgen receptors. In preclinical disease models, KBP-5074 has also shown better efficacy in lowering blood pressure and renal protection compared with the benchmark compound eplerenone. In addition, KBP-5074 was shown to be safe and well-tolerated when administered as ascending single doses (up to 30 mg) to healthy subjects and as multiple ascending doses in healthy subjects (up to 5 mg) and those with mild to moderate renal impairment (up to 2.5 mg) with minimal effects on serum potassium levels [data on file].

In order to further explore the potential use of KBP-5074 in subjects with advanced stages of CKD, the study described herein was designed to assess the safety, tolerability, pharmacokinetics (PK), and pharmacodynamics (PD) of a single dose of 0.5 mg KBP-5074 in subjects with severe renal impairment and those on hemodialysis (HD).

## 2 Methods

### 2.1 Study Design and Subjects

This clinical trial (ClinicalTrials.gov identifier: NCT02837237) was designed as a multicenter, open-label study in non-HD subjects (Part 1) and those on HD (Part 2) with severe CKD. Key eligibility criteria included male or female subjects aged 18 to 75, with a body mass index (BMI) between 19 and 42 kg/m^2^, who were nonsmokers or light smokers (smoked fewer than 10 cigarettes per day). Non-HD subjects in Part 1 had severe CKD defined as estimated glomerular filtration rate (eGFR) ≥15 mL/min/1.73 m^2^ and ≤29 mL/min/1.73 m^2^ based on the isotope dilution mass spectrometry traceable Modification of Diet in Renal Disease (MDRD) equation with serum potassium between 3.3 and 4.8 mmol/L, inclusive. Part 2 subjects were on a HD schedule for at least 45 days with KT/V ≥1.2 for end-stage renal disease (ESRD) with an average of 3 HD sessions per week.

In Part 1, a single cohort of 5 non-HD subjects with severe CKD received a single KBP-5074 dose of 0.5 mg after fasting between 2-4 hours. Following dosing, subjects were followed for 14 days for PK and safety assessments. These subjects remained confined to the clinic from Check-in (Day - 1) until 24 hours postdose and returned as outpatient visits for additional PK sampling up to 14 days.

In Part 2, a single cohort of 6 HD subjects received a single KBP-5074 dose of 0.5 mg on the same day of the first HD (immediately after the HD session). Following dosing, subjects were followed for 14 days for PK and safety assessments. The PK samples were obtained over 24 hours after dosing. Inlet (arterial/outlet (venous) PK samples were also collected during the second HD session and at the end of the second HD session. Venous plasma samples were collected following dialysis. The HD subjects remained confined to the clinic from Day - 1 until 52 hours postdose. Due to the long half-life of KBP-5074 (> 60 hours), it was necessary to assess the single dose PK over 312 hours to ascertain the cumulative impact of multiple HD sessions.

In both study parts, all subjects were followed for PK and safety assessments from dosing up through Day 14.

### 2.2 Pharmacokinetic and Pharmacodynamic Assessments

All blood samples were collected into tubes containing potassium ethylenediaminetetraacetic acid (K_2_EDTA). Samples were centrifuged at 1500 × g for 10 minutes at approximately 4°C within 30 minutes of collection. The resultant plasma was aliquoted and immediately frozen at −70°C or colder within 1 hour of collection.

In Part 1 (non-HD subjects), PK blood samples were collected at predose and postdose at 2, 4, 6, 8, 10, 12, 18, 24, 48, 72, 96, 120, 168, 216, 264, and 312 hours. Serum and plasma samples for biomarkers (aldosterone and serum potassium) were collected at predose and postdose at 2, 6, 12, 24, 48, 72, 96, 120, 168, 216, 264, and 312 hours.

In Part 2 (HD subjects), the first dose of KBP-5074 was administered immediately following a dialysis session. Additional dialysis sessions occurred at 48, 96, 168, 216, and 264 hours relative to the beginning of the first dialysis session. PK blood samples were collected immediately following the first HD session at predose and postdose at 2, 4, 6, 8, 10, 12, 18, and 24 hours. Samples were again collected during the second HD session (inlet [arterial]/outlet [venous]) at 44 hours postdose (immediately prior to HD at −5 min minutes and 5 minutes after the start of HD), at 44.25, 44.5, 46, and 47 hours postdose, and at the end of the second HD session. Following the end of the second dialysis session, venous plasma samples were collected postdose at 48.5, 49, 50, 52, 72, 96, 120, 168, 216, 264, and 312 hours. Cumulative dialysate was collected and weight/volume determined for the following intervals: 44 to 44.5 hours, 44.5 to 45 hours, 45 to 46 hours, 46 to 47 hours, and 47 to 48 hours postdose. Dialysate collection ended at the end of dialysis if dialysis was <4 hours.

Plasma samples and dialysate were analyzed for KBP-5074 levels using a validated analytical method using high performance liquid chromatography tandem mass spectroscopy (HPLC-MS/MS). The lower limit of quantification (LLOQ) was 0.1 ng/mL; the upper limit of quantification (ULOQ) was 50 ng/mL.

### 2.3 Safety Assessments

Safety assessments were conducted at predose and throughout the study and included the following: AEs, physical examination findings, clinical laboratory evaluations (chemistry, hematology, urinalysis), vital signs (blood pressure, pulse rate, respiratory rate, oral temperature), and 12-lead electrocardiograms (ECGs). These data were descriptively analyzed.

### 2.4 Statistical Analyses

The sample size for study parts was based on feasibility to provide sufficient data to characterize the PK of KBP-5074, without formal power calculations. A total of 12 subjects (6 subjects for each part) were planned for enrollment in the study. No formal statistical analysis of the safety data was performed. Plasma PK parameters were analyzed by non-compartmental methods and were descriptively analyzed. PK parameter calculations were based on actual sampling times relative to dose administration.

Log-transformed PK parameters (AUC_0-t_, AUC_0-∞_, and C_max_) were analyzed by analysis of variance (ANOVA). This analysis considered part (HD vs. non-HD) as a fixed effect. For each log-transformed PK parameter, the point estimate and its associated 90% CI were constructed for the difference between the HD group versus the non-HD group. Point and interval estimates of differences were exponentiated to obtain the ratio of geometric least squares (GLS) means and its 90% confidence interval (CI).

Hemodialysis extraction ratio (ER_D_) was calculated with the following equation: ER_D_ = (Ci-Co)/Ci, where Ci was inlet concentration (arterial) and Co was outlet concentration (venous). The estimated hemodialysis clearance (CL_D_) was calculated using this equation: CL_D_ = Q*(Ci-Co)/Ci, where Q was inlet flow. CL_D_ recovery was the amount recovered in dialysate/inlet AUC_0-t_. ER_D_ and CL_D_ were the average of the respective values across collection time points of 44 hours plus 5 min, 44.25, 44.5, 46, 47, and 48 hours postdose.

The PD variables (plasma aldosterone and serum potassium) were analyzed using a linear mixed model based on all observed data from Part 1 and Part 2. This model included change from baseline as response, part (HD vs non-HD), time point, and part by time point interaction as factors and baseline value as covariate, assuming a compound symmetric covariance structure among time points within each subject. Least squares (LS) means for each part, standard errors, associated 95% CIs, difference of LS means between parts, associated 95% CIs and nominal two-sided p-values were tabulated by time point. In addition, LS mean change from baseline and associated 95% CIs were plotted versus time point stratified by part for each of the PD variables.

## 3 Results

### 3.1 Demographics and Disposition

A total of 11 subjects (5 subjects with severe CKD in Part 1 and 6 HD subjects in Part 2) were enrolled in the study and all subjects completed the study. Other than the status of their CKD, the subjects were generally similar with respect to age, weight, and BMI (Table 1).

**Table 1.** Demographics and Baseline Characteristics

|  | **Part 1**<br>**Severe CKD**<br>**N = 5** | **Part 2**<br>**HD subjects**<br>**N = 6** |
| --- | --- | --- |
| **Age (yrs), mean (SD)** | 59.2 (14.62) | 48.7 (2.58) |
| **Gender, n (%)** |  |  |
| **Male** | 2 (40.0) | 4 (66.7) |
| **Female** | 3 (60.0) | 2 (33.3) |
| **Weight (kg), mean (SD)** | 88.6 (23.39) | 93.3 (16.40) |
| **BMI (kg/m<sup>2</sup>), mean (SD)** | 30.7 (4.72) | 32.0 (5.91) |

### 3.2 Pharmacokinetic Results

Mean plasma KBP-5074 concentrations peaked at 6 and 4 hours in non-HD and HD subjects, respectively, and slowly declined through 312 hours postdose (Figure 1). All non-HD subjects and the majority of HD subjects had measurable plasma concentrations at the last assessment time. Overall, the plasma KBP-5074 exposures were significantly lower in HD subjects compared with the non-HD subjects (Table 2, Table 3). The mean t_1/2_ was shorter in the HD subjects and the CL/F and Vz/F estimates were higher in HD subjects compared to non-HD subjects. As evident in Figure 1, HD did not directly alter the drug concentration. Inlet and outlet PK parameters during the second HD session are summarized in Table 4. During the HD, the AUCs for inlet and outlet samples were similar, indicating minimal impact of HD on drug concentrations. Only 2 subjects had values greater than 0 for ER_D_ and CL_D_ recovery: ER_D_ values of 0.0710 and 0.0898, respectively, and CL_D_ recovery values of 24.1 and 26.2 mL/min, respectively. KBP-5074 was below the level of quantification in the dialysate samples from all subjects.

**Figure 1.**
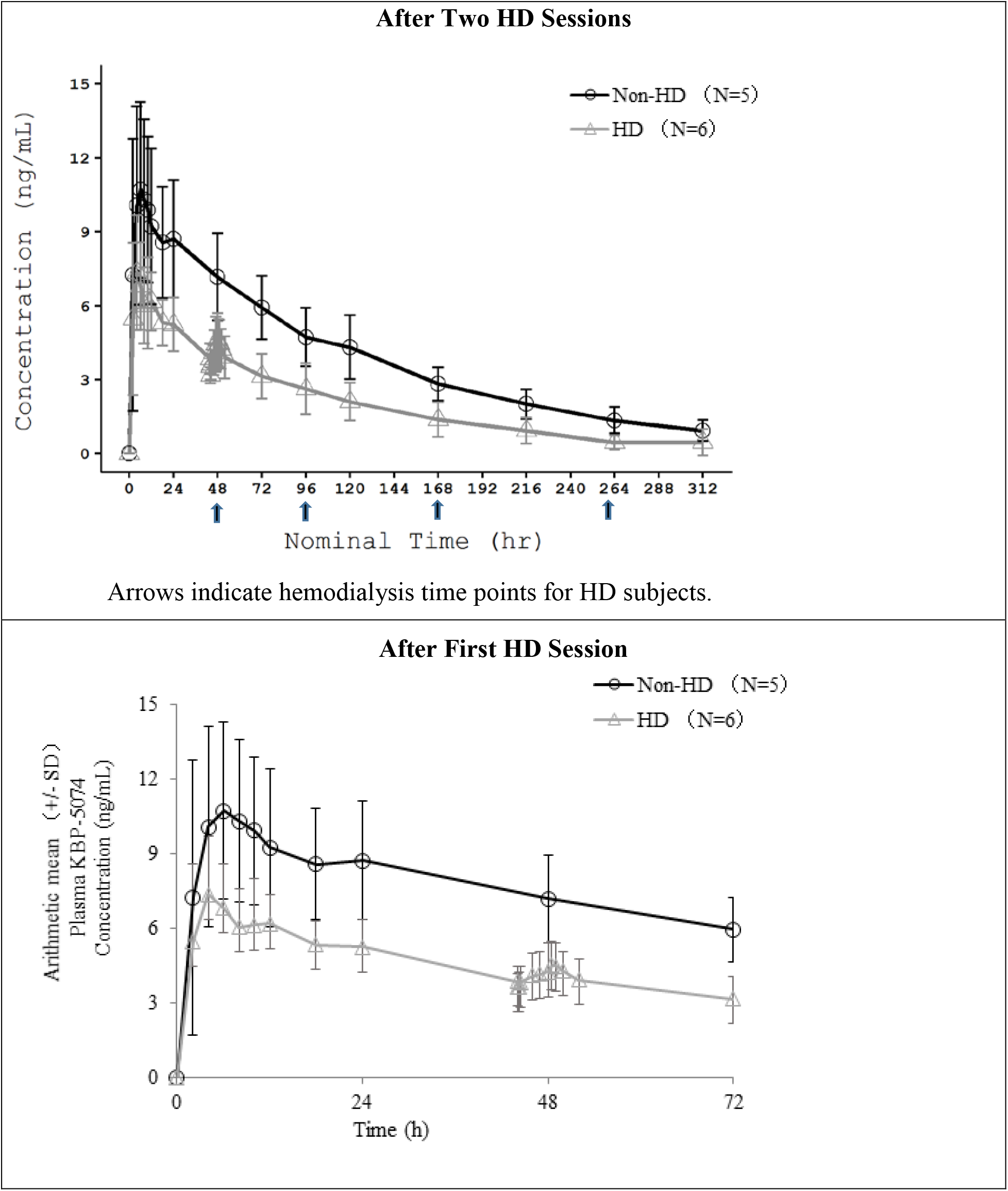
Mean (±SD) Plasma KBP-5074 Concentrations in HD and Non-HD Subjects

**Table 2.** Key Pharmacokinetic Parameters of KBP-5074 in Non-HD and HD Subjects Immediately Following the First Hemodialysis Session

| **C<sub>max</sub>**<br>**(ng/mL)** | **t<sub>max</sub>**<br>**(hr)** | **AUC<sub>0-∞</sub>**<br>**(hr\*ng/mL)** | **λ<sub>z</sub>**<br>**(1/hr)** | **t<sub>1/2</sub>**<br>**(hr)** | **CL/F**<br>**(L/hr)** | **V<sub>z</sub>/F**<br>**(L)** |
| --- | --- | --- | --- | --- | --- | --- |
| **Non-HD Subjects (N=5)** |  |  |  |  |  |  |
| 10.6<br>(38.7) | 4.00<br>(4.00, 6.00) | 1310<br>(24.0) | 0.00816<br>(29.3) | 84.9<br>(29.3) | 0.381<br>(24) | 46.7<br>(37.3) |
| **HD Subjects (N=6)** |  |  |  |  |  |  |
| 7.34<br>(28.1) | 5.00<br>(1.93, 12.00) | 669<br>(32.1) | 0.0111<br>(24.7) | 62.7<br>(24.7) | 0.747<br>(32.1) | 67.6<br>(16.2) |

Dialysis sessions occurred at 48, 96, 168, 216, and 264 hours following KBP-5074 administration.
Abbreviations: N: Number of subjects; C<sub>max</sub>: observed maximum plasma or serum concentration after administration; t<sub>max</sub>: time to reach the observed maximum (peak) concentration; AUC<sub>0-∞</sub>: area under the concentration-time curve from zero up to an infinite time; λ<sub>z</sub>: terminal elimination rate constant; t<sub>1/2</sub>: terminal half-life; CL/F: apparent clearance; V<sub>z</sub>/F: volume of distribution.
All data are presented as geometric mean (%CVb), except for t<sub>max</sub> for which median and range (minimum, maximum) are shown. %CVb = 100 \* SQRT [exp(S<sup>2</sup>)-1], where S is the standard deviation of the data on a log scale.

**Table 3.** Comparison of Pharmacokinetic Parameters for Non-HD and HD Subjects

| Parameter | Subjects | Geometric Mean | Ratio of Geometric Mean (%) | 90% CI for Ratio (%) |
| --- | --- | --- | --- | --- |
| **AUC<sub>0-t</sub>**<br>**(hr\*ng/mL)** | HD | 628 | 53.0 | 40.14 - 69.99 |
|  | Non-HD | 1190 |  |  |
| **AUC<sub>0-∞</sub>**<br>**(hr\*ng/mL)** | HD | 669 | 51.0 | 37.32 - 69.73 |
|  | Non-HD | 1310 |  |  |
| **C<sub>max</sub>**<br>**(ng/mL)** | HD | 7.34 | 69.3 | 48.39 - 99.11 |
|  | Non-HD | 10.6 |  |  |

N=5 for Non-HD subjects; N=6 for HD subjects
Ratio of geometric mean is calculated by HD/Non-HD (Test/Reference)
The ANOVA model included log transformed PK parameters as response and part as a fixed effect.

**Table 4.** Pharmacokinetic Parameters for HD Subjects during Hemodialysis Sessions

| Inlet<br>**AUC<sub>44-48h</sub>**<br>**(hr\*ng/mL)** | Outlet<br>**AUC<sub>44-48h</sub>**<br>**(hr\*ng/mL)** | Outlet:Inlet<br>**AUC Ratio** |
| --- | --- | --- |
| 15.5 | 15.8 | 1.02 |
| (20.0) | (20.6) | (7.13) |

Data are presented as geometric mean (%CVb).
%CVb = $100 * \text{SQRT} [\exp(S^2)-1]$ , where S is the standard deviation of the data on a log scale.

### 3.3 Pharmacodynamic Results

Plasma aldosterone and serum potassium concentrations were generally comparable between non-HD and HD subjects in Parts 1 and 2 (Figure 2). Aldosterone levels differed between the 2 subject populations at 6, 48, 216, and 312 hours postdose, with adjusted mean differences in the change from baseline (597.51 ng/L higher for non-HD subjects, 673.55 ng/L higher for HD subjects, 477.28 ng/L higher for HD subjects, and 381.35 ng/L higher for non-HD subjects, respectively). This is highly likely due to more variation observed in HD subjects than non-HD subjects.

**Figure 2.**
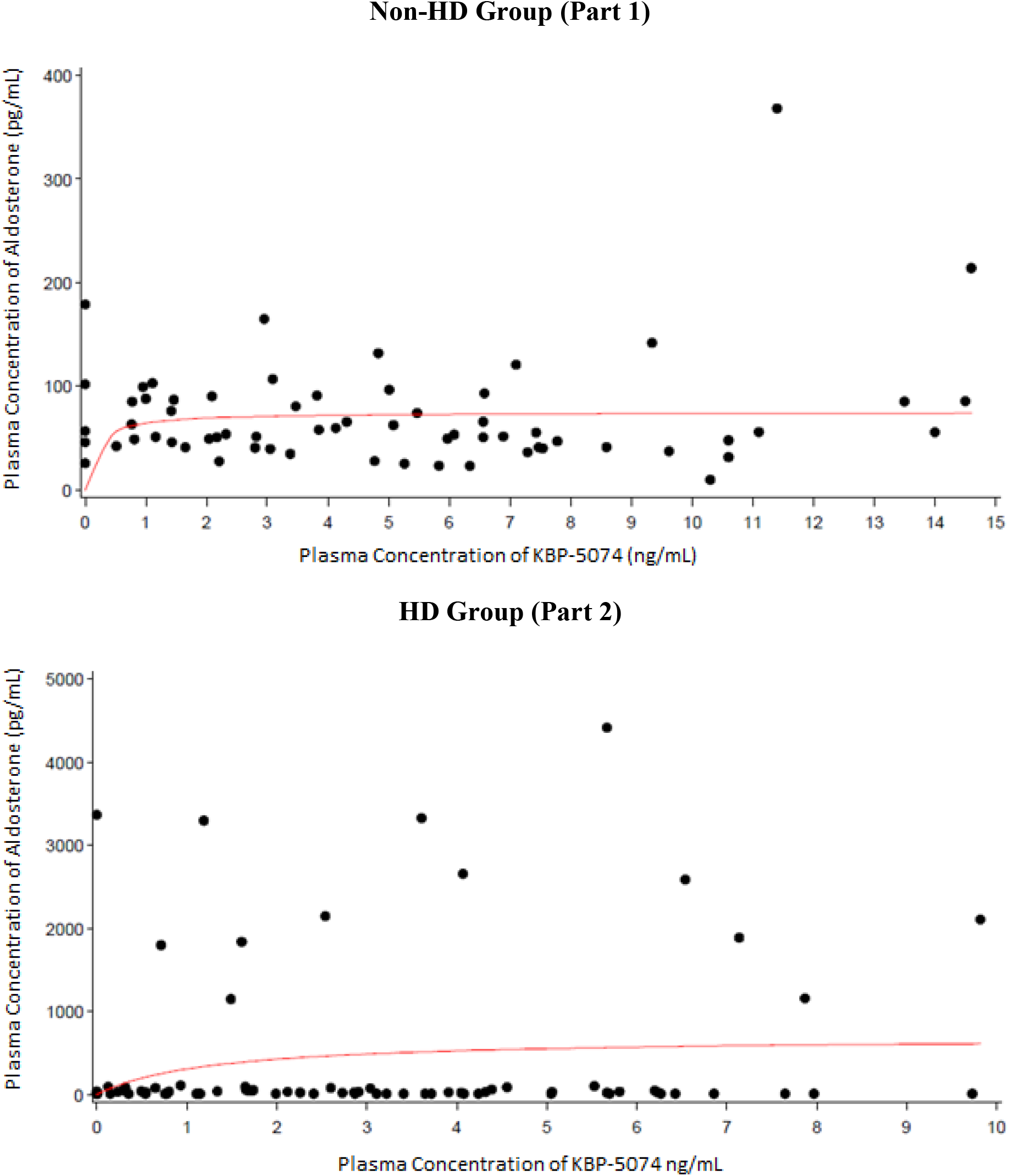
Plasma KBP-5074 Concentration vs Plasma Aldosterone Concentration

**Figure 3.**
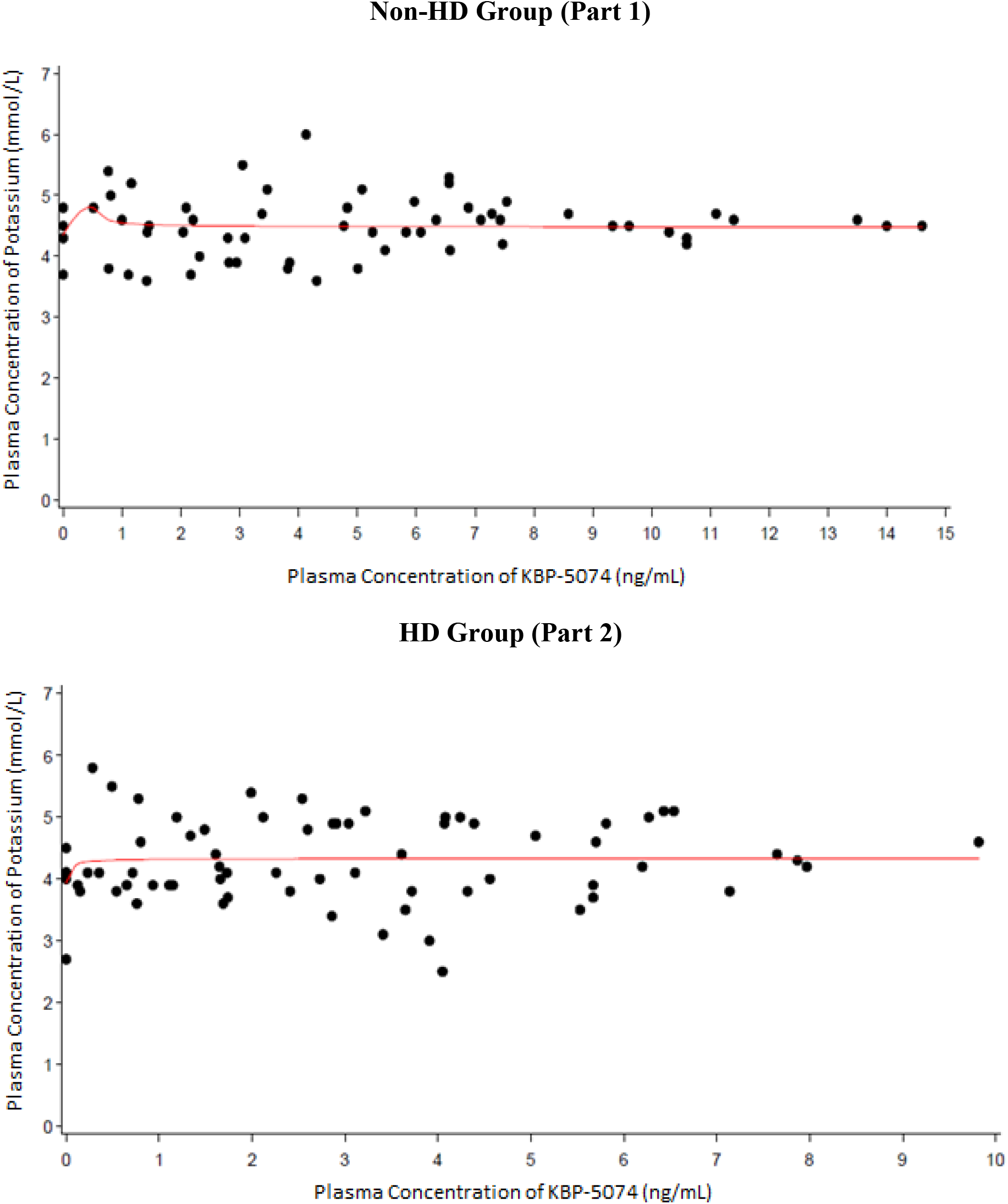
Plasma KBP-5074 Concentration vs Plasma Potassium Concentration

Potassium levels were comparable between non-HD and HD subjects, except some differences were observed at 48 and 72 hours postdose, with adjusted mean differences in the change from baseline (0.99 mmol/L and 0.66 mmol/L higher for non-HD subjects and HD subjects, respectively). These differences were highly likely due to greater variations observed in HD subjects.

A PK/PD analysis was conducted, but due to the small sample size, the results were not interpretable.

### 3.4 Safety Results

KBP-5074 was generally well tolerated by non-HD and HD subjects (Table 5). The number and frequency of AEs was similar between the 2 subject groups. The majority of AEs were mild in intensity with no reports of severe AEs. All AEs were considered drug-related, except for a single reported AE of hyperkalemia which was not considered drug-related by the investigator. During the study, one HD subject (Part 2) was reported with mild hyperkalemia on Day 14 days after receiving KBP5074 due to out-of-range potassium values, which returned to normal levels in a repeat test within 2 days. Overall, there were no clinically significant changes in laboratory values, ECGs, or vital signs. All AEs were reported as resolved by study end. There were no deaths or other serious AEs during the study.

**Table 5.** Summary of All Adverse Events

| **Preferred Terms** | **Non-HD Subjects**<br>**(N=5)** | **HD Subjects**<br>**(N=6)** |
| --- | --- | --- |
| Any AE, n (%) | 2 (40) | 2 (33) |
| Fatigue | 1 (20) | 0 |
| Procedural hypotension | 0 | 1 (17) |
| Blood creatinine increased | 1 (20) | 0 |
| Hyperkalemia | 0 | 1 (17) |
| Metabolic acidosis | 1 (20) | 0 |
| Headache | 1 (20) | 0 |

## 4 Discussion

MR inhibition has been shown to have a major role in interrupting the progression of kidney disease by considerably reducing proteinuria in patients with CKD.^2,3,4^ Currently, available treatments include the steroidal MRAs spironolactone and eplerenone that block aldosterone, but also bind to androgen, glucocorticoid, and progesterone receptors. The use of these agents is limited due to the potential risk for hyperkalemia, especially when administered along with inhibitors of the renin-angiotensin-aldosterone system (RAAS).^5^ Therefore, KBP-5074 is being developed as a new treatment option for hypertension and other diseases in advanced CKD patients. KBP-5074 is a novel non-steroidal MRA that preferentially binds the MR and inhibits the binding of aldosterone, a component of the RAAS. In two Phase 1 studies, administration of KBP-5074 was shown to be safe and well tolerated, with minimal effects on potassium, in healthy volunteers and in subjects with mild to moderate renal impairment [data on file]. The results from these studies provided informative PK and safety data and support further evaluation of KBP-5074 in ESRD in both HD or non-HD patients.

In the current study, the safety, tolerability, and PK of KBP-5074 were evaluated in subjects with end stage renal disease (ESRD). The results indicated that single dose administration of KBP-5074 at 0.5 mg was generally well tolerated in both non-HD and HD ESRD subjects. In the 11 subjects that received KBP-5074, there was a single report of mild hyperkalemia in one subject on Day 14 following drug administration. There were no other clinically significant findings during the study. Furthermore, there were no severe AEs, withdrawals due to AEs, deaths or other serious AEs.

The overall plasma exposure of KBP-5074 was statistically significantly lower in ESRD subjects on HD compared to non-HD ESRD subjects. The mean t_1/2_ was shorter in ESRD subjects on HD and the CL/F and Vz/F estimates were higher in ESRD subjects on HD compared to non-HD ESRD subjects. Despite an apparent lack of hemodialysis clearance of KBP-5074, exposures (based on C_max_ and AUC) of KBP-5074 in ESRD subjects on HD were consistently lower than observed in non-HD ESRD subjects. In this study, the exposures in ESRD subjects on HD (mean AUC_0-∞_ = 669 ng*hr/mL) was similar to exposures in subjects with normal renal function (mean AUC_0-∞_ = 678 ng*hr/mL) after a single dose of KBP-5074 0.5 mg [data on file]. In comparison, the exposure in non-HD ESRD subjects was higher at (mean AUC_0-∞_ = 1310 ng*hr/mL). The current study indicates that minimal KBP 5074 was cleared directly by HD. It is possible that lower exposures observed in HD subjects are due to secondary effects (e.g., impact on hepatic function) associated with HD in this population. While increased hyperkalemia was not observed in this study with renal impaired subjects, there might be a potential for increased hyperkalemia in ESRD subjects not on HD, due to higher exposures to KBP-5074.

## Data Availability

All data resides in the trial master file maintained by KBP Biosciences Co. Ltd.

